# Ethical and practical considerations in HIV drug trial closure: perspectives of research staff in Uganda

**DOI:** 10.1101/2020.11.14.20231720

**Authors:** Sylivia Nalubega, Karen Cox, Henry Mugerwa, Catrin Evans

**Affiliations:** Soroti University, Soroti, Uganda; University of Kent, Kent, United Kingdom; University of Nottingham, Nottingham, United Kingdom; Joint Clinical Research Center, Kampala, Uganda

**Author notes:** **Corresponding author** Sylivia Nalubega, Soroti University, School of Health Sciences, Department of Nursing, Uganda.

**Keywords:** HIV, trial closure, research staff, Uganda, ethics

## Abstract

There is a gap in evidence regarding how research trial closure processes are managed to ensure continuity of HIV care for HIV positive participants following trial closure within low income settings. This research aimed to establish how research staff in Uganda understood and practised post-trial care for HIV positive trial participants. A grounded theory study was conducted using in-depth individual interviews and focus group discussions with 22 research staff from 3 different trials in Uganda, in 2014-2015. The results indicated that researchers engaged in three main activities to support trial participants, including; (i) preparing for post-trial care, which included instituting trial closure guidelines, planning necessary resources, and informing trial participants about post-trial care; (ii) facilitating participants during trial exit by engaging in psychological and practical support activities, and (iii) providing follow up care and support for participants after trial exit, to respond to the needs of trial participants which often arose after trial exit. This study established a need for a holistic approach to post-trial-care of HIV positive trial participants in Uganda, and the need to engage multiple stakeholders including ethics authorities.

## BACKGROUND

Tackling the HIV epidemic has required an enormous, globally coordinated research effort. Much of this research has taken place in sub-Saharan Africa, due to the high prevalence/incidence of HIV, having readily available and willing volunteers, and the need to find suitable and affordable interventions for these settings (Selgelid & Jamrozik, 2018; Weigmann, 2015).

Research conducted in low income settings raises unique ethical concerns related to issues such as the low socio-economic status and low literacy levels of participants, and poor healthcare delivery in these settings, that makes them vulnerable to coercion (Weigmann, 2015). This has prompted advocates to call for a high level of ethical regulation of research conducted in these settings (Selgelid & Jamrozik, 2018). Currently most policy guidelines on trial conduct focus on issues around trial recruitment and implementation rather than closure. Recent research (by the authors) suggests that trial closure can be a stressful time for trial participants and that additional support may be required [Reference to our previous work].

Post-trial obligations have been largely understood to mean the obligation of researchers to provide a proven intervention to the respective trial participants beyond trial participation (Lawton et al., 2019). However, in some types of research, especially those involving chronic conditions, post-trial obligations may necessitate going beyond the provision of trial products/interventions, to incorporate a range of other on-going services such as continued care and management of the disease condition and related psychosocial services (Cho, Danis, & Grady, 2018; Lawton et al., 2019). In research involving HIV infected persons, there is need for continued/lifelong provision of HIV treatment, care and support, which requires referral and adequate linkage to alternative care facilities, and follow up beyond the period of trial closure (Cho et al., 2018). Additionally, the need for monitoring and compensation for potential adverse effects from trial interventions (Lawton et al., 2019), and provision of trial feedback (Chen et al., 2016; Schroter, Price, Malićki, Richards, & Clarke, 2019), have been key concerns in post-trial (research) ethics.

Despite a clear need, it is argued that the area of post-trial care is neglected both in practice and in research (Pratt et al., 2012; Slack, 2014). Areas such as recruitment, informed consent, standards of care during research, and monitoring and management of adverse effects during trial conduct receive greater attention than the issue of post-trial obligations (Nalubega & Evans, 2015). Existing research on post-trial care has been predominantly undertaken in clinical areas such as Cancer or Diabetes (Lawton et al., 2019). There is an important gap in our understanding of HIV-related post-trial practice.

### Study aim

This research sought to establish how research staff understand and practice post-trial care in drug trials involving HIV positive participants in Uganda.

## METHODS

The study adopted a qualitative constructive grounded theory approach (Charmaz, 2014), which resulted in construction of a model of Facilitated Transition reported in another paper.

### Study setting

We included participants from two research institutions involved in running three separate HIV clinical trials. Each of the trials was in a different geographical location. Trial 1 was conducted at an urban site in Kampala, Trial 2 at a peri-urban site in Western Uganda and Trial 3 at a peri-urban site in Eastern Uganda. Trials 1 and 3 were conducted by the same research institution.

### Recruitment and data collection methods

Purposive and convenience sampling approaches were employed to ensure representation of geographical sites, trial staff categories, and genders. In addition, the principle of theoretical saturation was adopted in determining the final sample size. We interviewed 22 research staff using either key informant interviews or focus group discussions. Research staff were approached through their supervisors, and all those approached agreed to participate. Staff were eligible to participate if they had worked directly with trial participants and had been involved in trial closure processes within the past one year.

Interviews were conducted in the English language by the first author (who was a Ugandan nurse and a PhD student at the time, who had previously worked on HIV clinical trials). All interviews were conducted from research clinics where the respective staff worked. Data was collected during October 2014 to August 2015.

### Data analysis

Interviews were transcribed and analysed using a standard grounded theory approach as described in Charmaz (2014) using open coding (line by line coding), focused coding (coding larger sections of data), axial coding (developing categories and showing their relationships between them), theoretical coding (comparing and collapsing categories), and theory construction. Other techniques that improved our analysis included memo writing, theoretical sampling, constant comparison, and diagramming. NVivo 10 was used to manage the data.

### Maintaining rigor

Measures to ensure rigor in the research process included constant discussions among the research team about the analysis and the resultant interpretations (Korstjens & Mosery, 2018), use of verbatim quotes to support our interpretations, and paying attention to disconfirming cases and opposing or divergent views of the participants.

### Ethics

Our study was approved by the University of Nottingham UK and The AIDS Support Organization (TASO) Uganda, Research Ethics Committee (REC). The study was registered with the Uganda National Council for Science and Technology (UNCST), as SS3608. We received written permission to undertake the study from both participating institutions and written informed consent was given by all respondents. Participants were assured of anonymity and total confidentiality of their information.

## FINDINGS

### Participants

The study included 22 research staff. These were: three trial coordinators, four clinicians, five counsellors/home visitors, and 10 nurses. Out of the 22 staff, 15 were from Trial 1, four were from Trial 2, and three were from Trial 3. The majority (72.7%) of staff were female. Supplementary file 1 provides details about the characteristics of included staff.

### Themes

The findings revealed a number of activities that researchers engaged in, or felt were necessary, to facilitate the transition of HIV positive trial participants from research to ‘usual care’ facilities for continued HIV management. These activities were interpreted into three major themes; (i) planning and preparing for trial closure, (ii) facilitating participants during trial exit, and, (iii) care and support after trial exit.

### Planning and preparing for trial closure

Researchers noted that providing post-trial care required prior preparation and, ideally, should be initiated with participants during preparation for the trial and during trial conduct. Researchers reported that they relied on ethical documents such as national and international research guidelines to prepare for post-trial care. However, they noted that most of these guidelines did not provide explicit guidance for post-trial care which made preparation difficult. For example, while areas such as informed consent and care during trial conduct were well elaborated in the guidelines, there was a lack of guidance on important post-trial issues such as follow up after trial closure to ensure adequate linkage to care or compensation for side effects related to trial participation. Consequently, researchers recommended the need for research guidelines to include post-trial care as an important part of trial practice.

> *If it is an obligation or if it is a policy of an institution, then they can add it (post-trial follow-up) on the budget; it can be added onto the budget and say ‘for us we do this, if it is a policy of an institution. (Jane, trial coordinator)*

Activities that were seen to be important to post-trial care included establishment of trial closure guidelines, planning necessary resources, and preparation of trial participants for post-trial care. Preparation of participants involved providing them with relevant information on trial closure and guidance on how and where to access care after leaving the trials. Psychological support was also provided to address the emotional needs of the participants associated with trial closure.

> *Trial closure starts at the beginning of the study apparently. Because as we start the treatment of patients, we go through the screening, we go through the enrolment, we see them settle in, we prepare them or we tell them that there is time when the study will end so that as we start, as they settle in, they know that there is time and the trial will end. So, by the time we get to the closure, they are already into closure. (Joy, counsellor/health visitor/community mobiliser)*

### Facilitating participants during trial exit

Trial exit was conceptualized as a phase during which trial participants are actively prepared for trial exit, their exit from the trial, and promoting linkage to a new HIV care facility. Research staff engaged in various activities to support these processes including continued psychological support to allay participants’ anxieties associated with leaving a trial.

> *Then of course, all the time we have to talk to the patients because some we know also they become a bit anxious, they have been with you for four years, may be for how many years, now somehow the end is coming, so you have to keep preparing them. (Jane, trial coordinator)*

Research staff reported the need to support trial participants to identify and link back to healthcare facilities of their choice. However, in practice, the support provided was generally limited to providing referral letters, an approach research staff perceived as passive and insufficient to achieve appropriate linkage to post-trial care.

> *Practically it ends at giving them referral letters, though usually on the referral letter we are giving, we have contacts, we put our contacts there as well in case the health giver the other side may need more information about what we have written. (Lydia, clinician)*

Research staff felt that current practices regarding linkage to care could be improved by engaging in a more proactive researcher-led process, for example, by accompanying participants to the care facilities and assisting them to (re)register in care.

> *For example, these patients we work with, some have challenges that we don’t write in the exit reports, for example a patient is having psychosocial issues, a patient is having adherence issues, a patient is having may be some health issues or medical issues, that would be discussed doctor to doctor. So I believe it would be good when we move, we see those health workers, we discuss with them on the way forward of the patient other than giving them exit reports that don’t explain more” (Favour, counsellor/home visitor)*.

In addition, due to the impoverished situation of many trial participants, staff strongly felt the need for some continued financial/material support to address the socio-economic needs of the participants after trial closure. This need was partly attributed to an ethical obligation of researchers to compensate trial participants for their contribution in the trials and partly seen as a moral obligation to support those who were in need and who had become accustomed to receiving benefits during the trial.

> *So me my appeal to researchers is to always at the end of the study to extend some help to those people, because they give in a lot. (Favour, counsellor/home visitor)*

### Care and support after trial exit

Upon leaving the trial, research staff reported the need for participants to be supported as they established themselves back into ‘usual’ health and care routines. At a minimum, staff felt that participants should be supported for a period of 12 months, during which a number of activities should be undertaken to provide psychological and socioeconomic support to enable participants to adjust to life after the trial.

> *“It makes a lot of sense to follow them up because for some drugs, the reactions or side effects may come a little later than within the defined study period. So it is important to follow them up and see if anything came up that would still be associated with the drugs, but it is not done. At least we don’t do it as an institution”. (Wambo, clinician)*

In addition, all staff saw it as their duty to provide feedback of trial outcomes. Despite this desire however, dissemination of trial results had not been done in any of the included trials. Staff cited bureaucratic reasons, such as trial regulatory issues, as sometimes interfering with timely delivery of trial results. They recommended that a mechanism should be provided in which participants can receive interim results as they wait for the final trial feedback.

> *… you know these regulatory issues, because sometimes it depends on which scientific conference we are going to present. So you cannot disseminate results before the scientific conference has…(Destiny, nurse)*

Additionally, accessing participants for trial dissemination after trial exit was reportedly difficult due to changes of contacts or relocations by participants.

Research staff reported that despite their willingness to offer support after trial exit, it was practically difficult. This was because post-trial care activities are usually not planned for, and hence were not budgeted for. Without the requisite resources, no follow up could take place.

> *The reason as to why it (post-trial follow-up) is not done is because the sponsors facilitate the study up to the date of exit, we stop there. (Favour, Counsellor/home visitor)*

Overall, the support provided to trial participants after trial exit was minimal, and relied upon the individual initiatives of the staff. It was strongly suggested that the implementation of post-trial care would require a collaborative approach between a range of stakeholders. Stakeholders would include the researchers, health facility workers, local NGOs, the community, the Government, and ethical bodies. These would play key roles in streamlining the provision of post-trial care, from instituting policies to actual provision of care and monitoring of participants until they settle into post-trial care facilities.

> *So bringing people on board where we are referring is also important, which has not been there, we do plan other things, we do plan the end of trial as researchers this side, and it is only at the time of exit that we do give them this letter as an introduction, these people are unaware of what else has been happening, we really need to put these people on board before closure. (Alloy, trial coordinator/nurse)*

## DISCUSSION

This study aimed to establish how research staff understand and respond to the needs of HIV positive trial participants during closure of the trials. Researchers reported a number of activities that are needed to address the needs of the trial participants and to facilitate their smooth transition from research to ‘usual’ care facilities. To prepare participants for trial exit, researchers placed great importance on the role of counselling and emotional support in addressing the post-trial care needs of the participants. This approach was considered important to reduce negative emotional effects associated with trial closure (Lawton et al., 2019), and to offer guidance to participants on the next steps in accessing HIV care.

Continuity of care after trial closure requires that there are appropriate processes in place to support trial participants to be linked to alternative facilities (Odero et al., 2018). In HIV and other chronic disease research, appropriate linkage to care is important, given the potential negative consequences of treatment interruptions or treatment failure (Odero et al., 2018). Participants in the current study reported using referral for linking participants to post-trial care which they considered passive and unreliable in facilitating successful linkage. Although established international research institutions (Rennie & Sugarman, 2009; UNAIDS, 2012; UNCST, 2014) consider referral as an acceptable approach to post-trial care, this approach has been criticised by various authors who confirm this study’s finding that a more practical, proactive and staff-facilitated approach to HIV care linkage is required (Koduah Owusu, Adu-Gyamfi, & Ahmed, 2019). A more proactive approach has been successfully used to link HIV positive individuals to care following HIV testing and research has demonstrated higher rates of linkage to, and retention in, HIV care (Elul et al., 2017). Such an approach could be adapted for HIV trial closure. Currently, data on linkage to care following research participation is lacking (Koduah Owusu et al., 2019) and this is an important research gap. The possibility of negative side effects occurring after trial closure was a major concern for researchers in this study. Many cited a need for on-going monitoring of trial participants for some time following trial exit. The same concern has been expressed by several other authors (Lawton et al. (2019), and Bukenya, Seeley, Tumwekwase, Kabunga, and Ruzagira (2020)) who demonstrate that additional follow up and support of HIV positive clients following linkage to care significantly improved their general outcomes. The need to compensate trial participants for their engagement in research was another issue raised in the current study, and is also supported in literature (Kwagala, Wassenaar, & Ecuru, 2010). However, the whole area of compensation after a trial remains under researched with knowledge gaps to understand the amounts and impacts of financial benefits among resource constrained individuals.

Finally, dissemination of trial results to participants was cited as an important researchers’ responsibility. However, although an important part of clinical trials, some authors have reported that most volunteers actually never receive trial feedback (Chen et al., 2016; Schroter et al., 2019). In the current study, hindrances to dissemination reported included trial regulatory issues and challenges in accessing participants once they leave the trials. These findings indicate the need for early dissemination of trial results and for provision of interim results where possible, before participants area exited. Moreover, limited documented evidence exits on the practice of trial feedback in HIV research which calls for more research.

Our study showed that post-trial follow up and monitoring was rarely achieved. It is advised to have plans for follow up and monitoring incorporated in the entire research protocol (Odero et al., 2018). Moreover, to enable standardisation of post-trial care among researchers, post-trial care guidelines should be incorporated in research ethics policies and enforced by the ethical authorities (Lawton et al., 2019; Pratt, Paul, Hyder, & Ali, 2017). This study also identified a need for stakeholder involvement in HIV post-trial care. Various stakeholders including health facilities, local leaders and NGOs could be essential in managing trial participants after they leave research related care (Tso et al., 2016).

## Conclusion

This study aimed to establish how research staff understood, experienced and practiced post-trial care for HIV positive trial participants in Uganda. The findings revealed a number of activities that researchers engaged in or felt were necessary to facilitate the transition of HIV positive trial participants from research to usual care facilities for continued HIV management. In addition to ensuring continued access to trial medications (through referral back to routine care) and providing trial feedback, the research identified other critical needs to be incorporated in post-trial care, including; follow-up care and monitoring, and financial support. There was general recognition for the need to involve various stakeholders at different points of the transition process, and ethics authorities were viewed as important actors in the implementation of post-trial care, by their role in instituting and enforcing post-trial care policies.

## Data Availability

All data refered to in this manuscript can be accessed with permission from the corresponding author

## Funding statement

This research received no specific grant from any funding agency in the public, commercial, or not-for-profit sectors. The Ph.D programme under which the study was undertaken was sponsored by the Vice-Chancellor’s Scholarship for Research Excellence (International) and the University of Nottingham, United Kingdom.

## Conflict of interest disclosure

The Authors declare that there is no conflict of interest.

## Notes

### Competing Interest Statement

The authors have declared no competing interest.

### Author Declarations

Ethical approval was received from the University of Nottingham, UK and The AIDS Support Organization (TASO) Uganda, Research Ethics Committee (REC). The study was registered with the Uganda National Council for Science and Technology (UNCST), as SS3608. Permission to conduct the research was granted by the respective research institutions, and written informed consent was received from all respondents.

